# Importation Risk Stratification for COVID19 using Quantitative Serology

**DOI:** 10.1101/2021.09.29.21264323

**Authors:** David E Williams

## Abstract

Recent work (Khoury *et al*.,*Nature Medicine* **2021**, *27* (7), 1205-1211) has shown that measurement of IgG antibody concentration in blood correlates well with vaccine efficacy. The present communication builds on this work and considers the probability of infection given immunity, taking into account the distribution across the population of antibody concentration in vaccinated or convalescent people. The model is consistent with the observed rates of breakthrough infection following vaccination or previous infection. The model is then developed to consider the use of quantitative measurement of antibody concentration on arrival as an aid to risk stratification of travellers. The model indicates that such a measurement could significantly decrease the quarantine time required to achieve a given level of importation risk.

## Introduction

This communication considers importation risk of COVID from travellers. In particular, it considers the use of quantitative measurement of COVID antibody concentration as part of a risk assessment framework applied to incoming travellers. Currently, countries are deploying different levels of control on entering travellers, based on COVID prevalence and risk appetite in both arrival and departure country. Source country risk stratification, pre-departure testing based on either symptoms or virus testing or both, with different testing regimes, and varying quarantine requirements with or without viral testing are being used. Tests may be on saliva or from naso- pharangeal swabs, and may be for viral antigen or for viral RNA. The objective of the control regime is to reduce to an acceptable level the risk of importation of COVID, which would vary from one country to another. Figure 1 illustrates a control regime and notes the inputs required in order to model the risk that an infected traveller might be released into the community.

**Figure 1.**
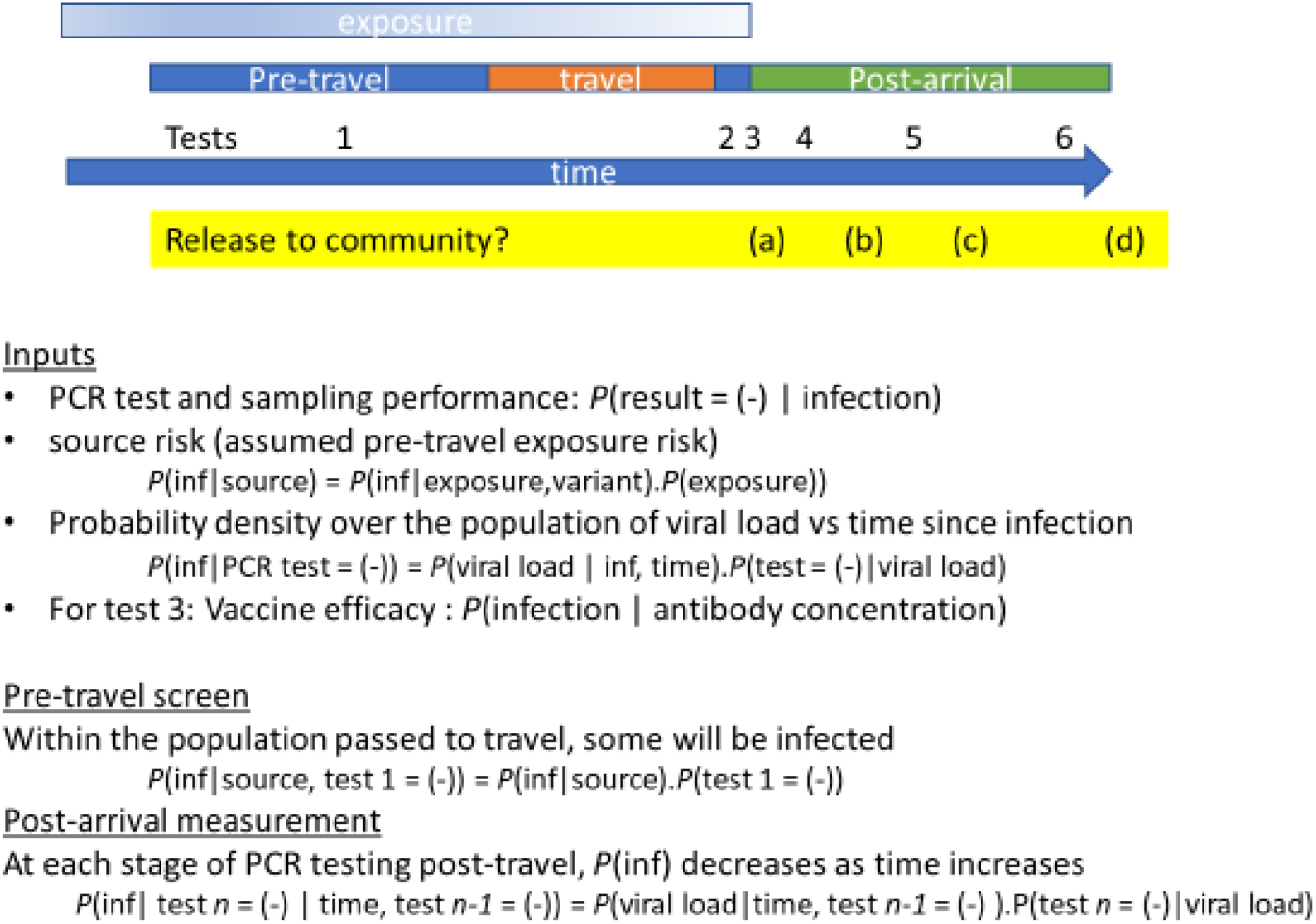
Illustrative border travel control procedure. In this model, a quantitative antibody test is introduced as test 3.

Modelling risk of releasing an infected traveller into the community requires knowledge of a number of conditional probabilities: for example the probability of a false negative virus test result *P*(result = (-) | infection). That probability depends on the viral load the person is carrying, how the sample is taken (probability that the sampling method and device will capture virus) the analytical sensitivity of the test (limit of detection) and the intrinsic variability of the analytical result. The viral load varies significantly from one person to another and strongly with time since the initial infection ^1, 2^. Yang *et al*^3^. and Smith *et al*.^4^ have recently described models for importation risk assuming various controls and different source prevalence. Given knowledge of the probability of infection given exposure that may be derived from population-scale studies of infection ^5^, knowledge of test performance in practise derived from repeat testing, an assumed time course of viral load following infection, and an assumed model for the distribution across the population of parameters of the viral load, “travellers” are sampled at random and followed through the steps in the testing regime to evaluate how many infected ones are undetected. Yang *et al*.^3^ assumed pre-departure PCR testing, symptom observation and on-arrival PCR testing followed by different quarantine and test regimes. Smith *et al*. additionally considered the effect of classifying source countries according to disease prevalence as a way of modifying quarantine requirements ^4^.

Given the roll-out of vaccination, there is much current discussion concerning the use of vaccination certificates as an additional control that might shorten or avoid quarantine requirements: an additional control on arrival that is represented as ‘test 3’ in Figure 1. One consideration therefore is how to verify vaccination status, and also significantly reduce reliance on digital or paper-based vaccination verification methods that have high potential of being inaccurate, inconsistent between countries, and potentially insecure. Serology (detection of antibodies in blood) has been trialed in at least one jurisdiction ^6^. Furthermore, although vaccines are very effective in preventing infection, vaccination break-through infections particularly associated with the Delta variant and dependent on the vaccine type are well-documented ^5, 7, 8^. So, there has been discussion on how a simple serology result may be combined with classification by origin of travel together with both quarantine and PCR testing in order to shorten quarantine whilst also keeping risk to an acceptable level ^6^. Antibody concentration in response to vaccines varies significantly from one individual to another, and may to some degree be classified according to age and gender for example ^9^. Neutralising antibody concentration correlates with vaccine efficacy ^10-12^ and appears to decrease with time following vaccination. The risk of breakthrough infection appears to increase with time following vaccination ^13^. Quantification of antibody concentration of an individual therefore would appear to be an important element for assessing risk associated with an individual traveller. Therefore, this document considers the additional reduction of risk that may be obtained by employing a quantitative immunity test on arrival.

### Problem statement

The problem is framed in terms of conditional probabilities. There are two parts: probability that a person is infected and infectious given some assessment, and probability of infecting others. Probability of infecting others depends on viral load, variant and contact (behaviour) as well as on the proportion of the receiving population that is vaccinated. It has been demonstrated that “whilst vaccination may reduce an individual’s overall risk of becoming infected, once they are infected there is limited difference in viral load (and Ct values) between those who are vaccinated and unvaccinated.”^14^ Whilst the probability of infecting others can be managed by controls, conservatively we assume that, if a person is infected then they will be infectious and transmission to others will occur, dependent only on the proportion of the receiving population that is vaccinated. Hence we wish to assess the probability that an individual is infected: *P(infected)*

*P(infected)* depends on immunity (antibody concentration), variant and contact with other infected people, which can be expressed as a probability of transmission, *P(transmission)*

*P(transmission)* depends on variant, disease prevalence in source population, vaccination status of the receiving population and probability of contact (behavioural variables, societal control measures). The conditional probability model is

*P(transmission) = P(transmission*|*exposure)*.*P(exposure)*

Probability of transmission given exposure, *P(transmission*|*exposure)* depends on the variant and the nature of contact (close, casual) as well as on the vaccination status of the receiving population.

So :

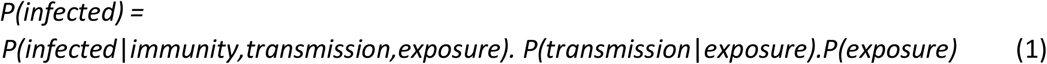

The problem is then to derive estimates for each of these terms, leading to an estimate of a threshold, *T*, such that *P(infected)* is less than some desired level. A model which incorporates additional controls, such as those given by Yang et al.^3^ and Smith et al.^4^ can be considered in part as an estimate of the product *P(transmission*|*exposure)*.*P(exposure)*.

### Model development

The problem is first to obtain an estimate *P(infected*|*immunity)*. We assume a relationship between vaccine efficacy and neutralising antibody concentration^10^. Specifically, we wish to determine for any given individual presenting for assessment the probability *P(infected*|*immunity,T)* where *T* denotes some given antibody threshold concentration. For the purpose of assessing the model, we also wish to determine the infection risk in a vaccinated population – the population breakthrough infection risk. For the purpose of using the model to evaluate an incoming population, we would wish to evaluate the incoming breakthrough risk.

The starting point is overall vaccine efficacy (VE) in clinical trials, estimated from the infection rates amongst vaccinated and unvaccinated populations: VE = 1- ARV/ARU where ARV is the attack rate in vaccinated people and ARU is that in unvaccinated people. ARV = (number of vaccinated people who become infected / total number of vaccinated people), where the *P(exposure)* and *P(transmission*|*exposure)*.is assumed the same for both vaccinated and unvaccinated groups over the time period of assessment. Vaccine efficacy in a population study is therefore derived from the observed population infection rate. Khouri *et al*.^10^ correlated VE with antibody (IgG) concentration in blood measured as part of the same trials. Cromer *et al*. showed correlation between these originally measured IgG concentrations and neutralising antibody concentration determined in live virus assays^15^, in order to deduce levels of immunity appropriate for different virus variants. Input data are the estimated dependence of vaccine efficacy for some antibody concentration, *n* : *E*_*I*_*(n)*; and the antibody concentration distribution across the vaccinated population, *f*_*V*_*(n)*.

An issue with literature antibody concentration data is the lack of an agreed scale. Data are variously reported as titre, as arbitrary units dependent on the assay, as IU scaled to a WHO standard pooled convalescent sample from 2020^16^, or as ng mL^-1^ based on a standard antibody. The results depend on the specific antigen used in the assay: whether whole spike protein (S), binding subunit (S1) or receptor binding domain (RBD); and on the particular antibodies or antibody population measured. Khouri *et al*.^10^ approached this problem by assuming that the distribution of antibody concentration in a convalescent population was invariant across different populations, and that it could be adequately described as a log-normal distribution with mean *μ*_*c*_ and standard deviation *σ*_*c*_. Because all the studies used different concentration scales, they expressed concentration as a ratio to the geometric mean (mean of the log-normal distribution) of the concentration distribution for convalescent patients, measured in the same study. For some of the studies, the number of samples was small so estimates are uncertain. Figure 2 shows concentration distributions for convalescent patients and for two different vaccines, based on the values for log-normal mean and standard deviation given by Khouri *et al*^10^. Here, the log-normal standard deviation for all groups is assumed to be the same as that for the convalescent patients, for which the value was more reliably established. The combined impact of prior infection and vaccination on antibody concentration distribution is to shift the mean to much higher values ^17^ which, given a log-normal distribution, has the effect of further broadening the distribution.

**Figure 2.**
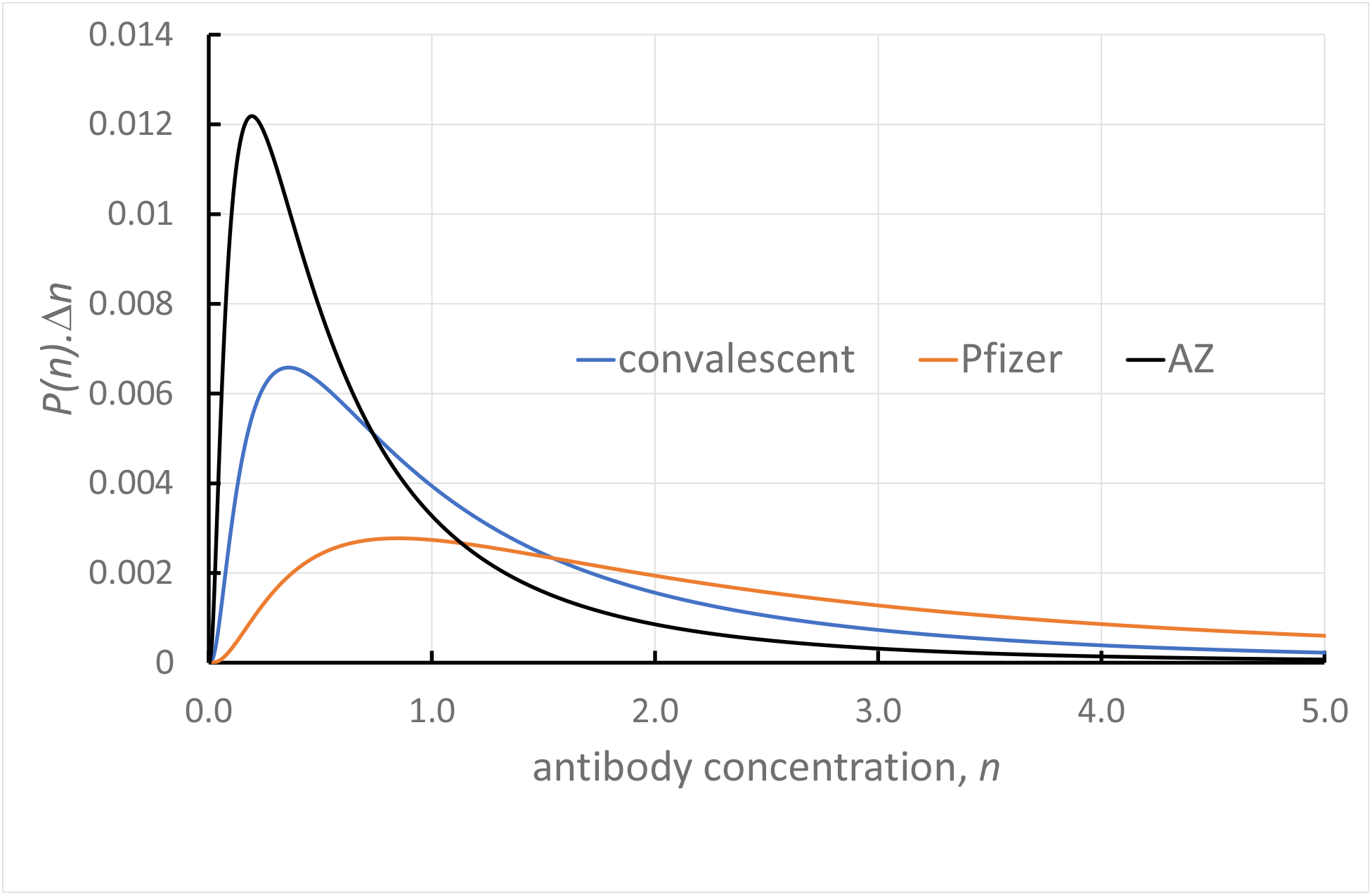
Antibody concentration distributions (log-normal) deduced for convalescent individuals and those vaccinated with Pfizer and AstraZenica vaccines. Concentration, n, relative to geometric (log-normal) mean convalescent concentration.

The model of Khouri et al.^10^ for vaccine efficacy as a function of neutralising antibody concentration is:

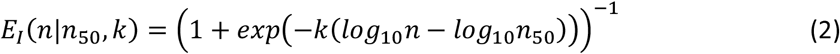

Where *E*_*I*_ denotes the vaccine efficacy at antibody concentration, *n*, and *k* and *n*_*50*_ are parameters fitted to vaccine efficacy data. Figure 3 shows the result of this model.

**Figure 3.**
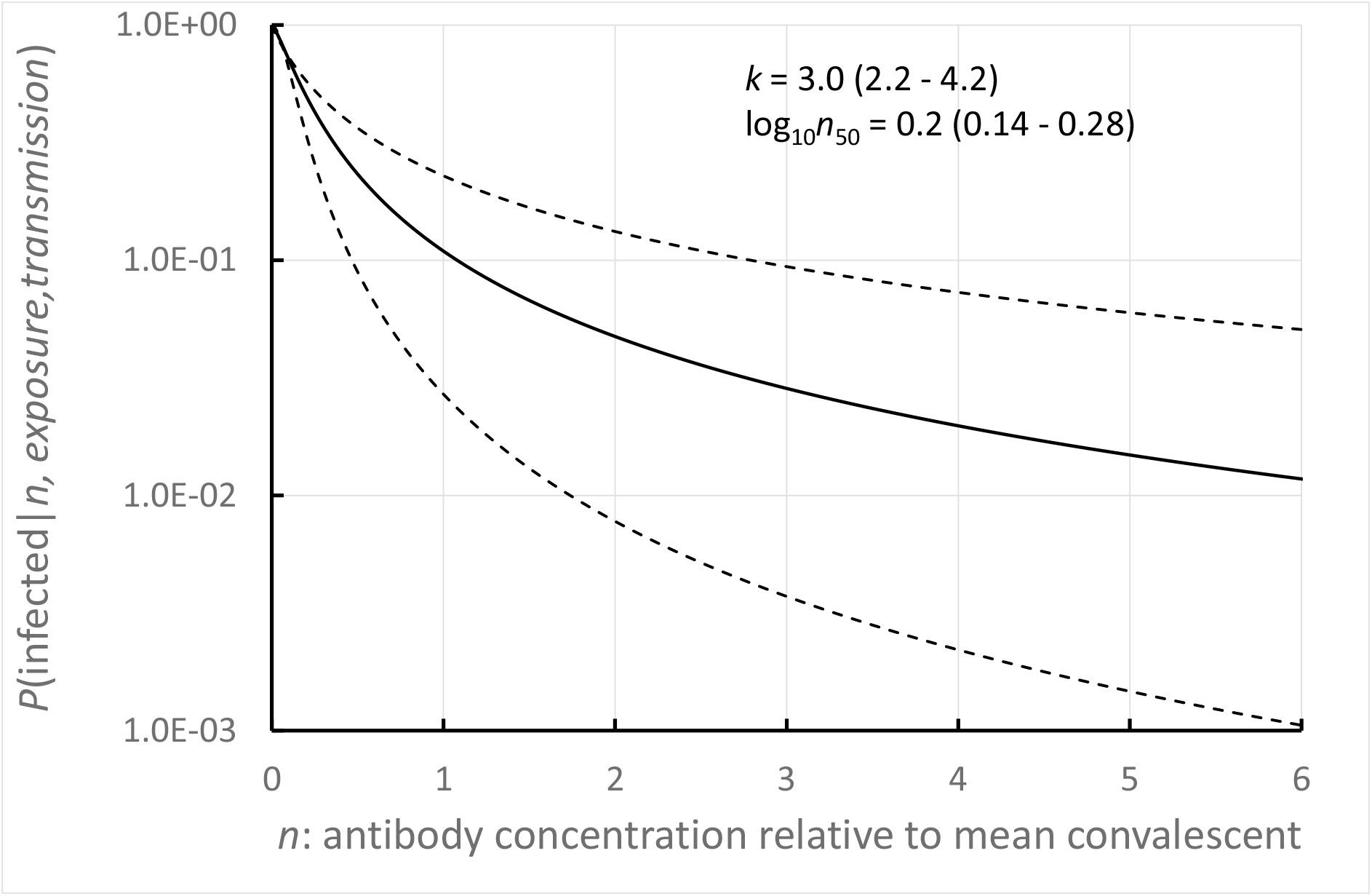
Individual risk of being infected given antibody concentration n, given exposure and transmission. Model of Khouri et al. with their parameter estimates (95% confidence intervals). Solid line gives the most likely estimate. Dotted lines give extreme estimates (upper limit k with lower limit n_50_, and lower limit k with upper limit n_50_).

The significance of the model embodied in equation 2 is that *E*_*I*_*(n)* is not dependent on the type of vaccine; nor is it dependent on the details of the antibody concentration distribution across the population. Use in a model for assessing individual risk does not depend on any assumption regarding the antibody concentration distribution, other than the assumptions used to construct the model. Use in a model to assess population risk does require an assumption about the population antibody concentration.

#### Population risk

Population risk is the risk of infection across an entire population, given exposure and transmission, taking into account the antibody concentration distribution across the population:

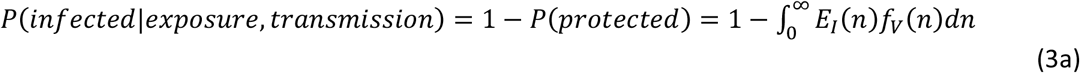

Across a population, *f*_*V*_*(n)* would depend on the proportion of vaccinated vs unvaccinated people, proportion of different vaccines used and the antibody concentration distribution for each individual vaccine, the proportion of previously infected people and whether previously infected people have been vaccinated or not ^9, 11, 12, 17-24^.

#### Individual risk based on an individual measurement, n

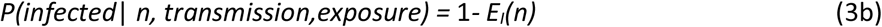

#### Incoming individual risk based on a threshold, T, applied to each individual measurement

This scenario applies for example where a measurement is used to determine ‘vaccinated or not’ with a decision based on whether the result is above some threshold. As the threshold increases, the proportion of the population who fall above the threshold decreases so the probability that any one person sampled from the whole population may be infected decreases. *P(protected)* in eq (3a) is normalised by the proportion of the population having concentration above the threshold:

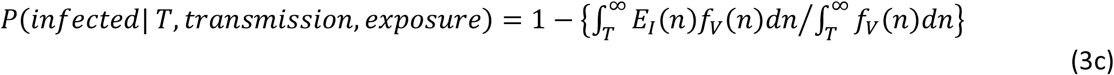

#### Model validation

The model can be validated, at least to some degree, by comparison of predictions with observed breakthrough infection rates, determined across a population: that is, by application of equations (1) and (3a)

##### (a)Assessment of probability of transmission (being infected) given exposure

For the assessment of *P(transmission*|*exposure)* data from Public Health England on secondary attack rates are used^5, 14^: known cases where the nature of the contact giving rise to the infection could be established. The data are for the probability of infection. Since these are therefore known infections, *P(exposure)* = 1 so *P(infection)* = *P(transmission*|*exposure)*. The probability of transmission given exposure depends on the variant and the nature of contacts. “Contacts” in the datasets is a binary variable : ‘household’ (‘close’); or ‘non-household’ (‘casual)’. These numbers are population averages and not values for any given individual and as such are an approximation. Table 1 gives the numbers.

**Table 1.** Probability of transmission (secondary attack rate) by variant and contact ^5, 14^ taken as. P(transmission|exposure). Secondary attack rates were based on positive tests amongst named contacts by an original case identified with a confirmed or probable variant.

|  | Non-household | household |
| --- | --- | --- |
| alpha | 0.042 | 0.096 |
| delta | 0.061 | 0.111 |

##### (b)Assessment of probability of exposure

Assessment of *P(exposure)* depends on a number of factors^3^, specifically the prevalence in the source population but also behavioural factors such as age and gender and address (rural / urban). Based simply on prevalence in the source population WHO data^25^ shows, *P(exposure) ≈ reported cases in previous 7 days / population* would vary from ∼ 0.1 to less than 1 × 10^-6^. For the purpose of modelling, Smith *et al*.^4^ assumed three probability classes, grouping source countries as 0-1, 1-10, and 10-100 prevalence / 100,000 population.

##### (c)Effect of variant

For the delta variant, the higher probability of transmission via contact, to unvaccinated people, is known and the vaccine effectiveness lower^8, 26, 27^. Wall *et al*. ^11, 12^ have considered the effect of variant on the model given by Khouri *et al*. and concluded that results of decreased activity in a live-virus neutralisation assay could be explained by a decrease over time in population antibody concentration. They considered that the model of Khouri *et al*.^10^ remained valid: that is, for estimates of risk, equation (2) remained valid with unaltered parameters so the effect of the variant type could be accommodated solely in the transmission probability. That assumption is retained here. However, there is developing evidence that the population of antibodies induced by infection and by current vaccination may be less effective against the delta variant^26, 28, 29^. The effect would be expected to be observed in the fitting parameter *n*_*50*_ (equation 3). Indeed, Cromer *et al*.^*15*^ deduced that lower vaccine effectiveness against the delta variant was due to a reduced proportion of effective antibodies within the antibody response to vaccination, and showed that the model could be appropriately adjusted by shifting the antibody concentration scale.

## Results

### 1. Assessment of model for population risk using known vaccination breakthrough rates

Assessment can be made for cases where a reasonable assumption is *P*(*exposure*) = 1. This is for health-care workers. Two datasets are available: the SIREN study of healthcare workers in England^5^, and a study from Israel, of breakthrough rates in healthcare workers vaccinated with the Pfizer-BnT vaccine^7^. The SIREN cohort is > 95% fully vaccinated; some are COVID-recovered; people in the UK have been vaccinated with the both the Pfizer vaccine and the AstraZenica vaccine. Breakthrough infections in in England are now dominantly of the delta variant. For the study by Bergwerk et al.^7^, 85% of the breakthrough infections were of the alpha variant. Another assessment can be made using data for known re-infection rates, given by Public Health England, which are also classified by variant type^5^. The data give the fraction of all infections within the assessment period that were identified as re-infections. Thus, given that these are all known infections, *P*(*exposure*) = 1. Equation (3a) was used for the calculation with *E*_*I*_*(n)* given by equation 2 with the most probable parameter values given by Khouri et al. Antibody concentration distributions shown in Figure 1 were used in the calculations. For the re-infection cases, a reasonable assumption is that these were dominantly due to due to non-household contacts. For this class, the convalescent antibody concentration distribution was used. For health-care workers, the risk of transmission would be expected to be higher, so transmission risk for household (close) contacts is used in the modelling. For this result, for the SIREN study given uncertainty about the proportion of the different vaccines used, again the convalescent antibody concentration distribution was assumed. For the study by Bergwerk et al.^7^, the antibody concentration distribution used was that for the Pfizer vaccine and the transmission probability was weighted to reflect the relative proportion of the alpha and delta variants. Figure 5 shows the result. Agreement is reasonable, given the assumptions and approximations.

**Figure 5.**
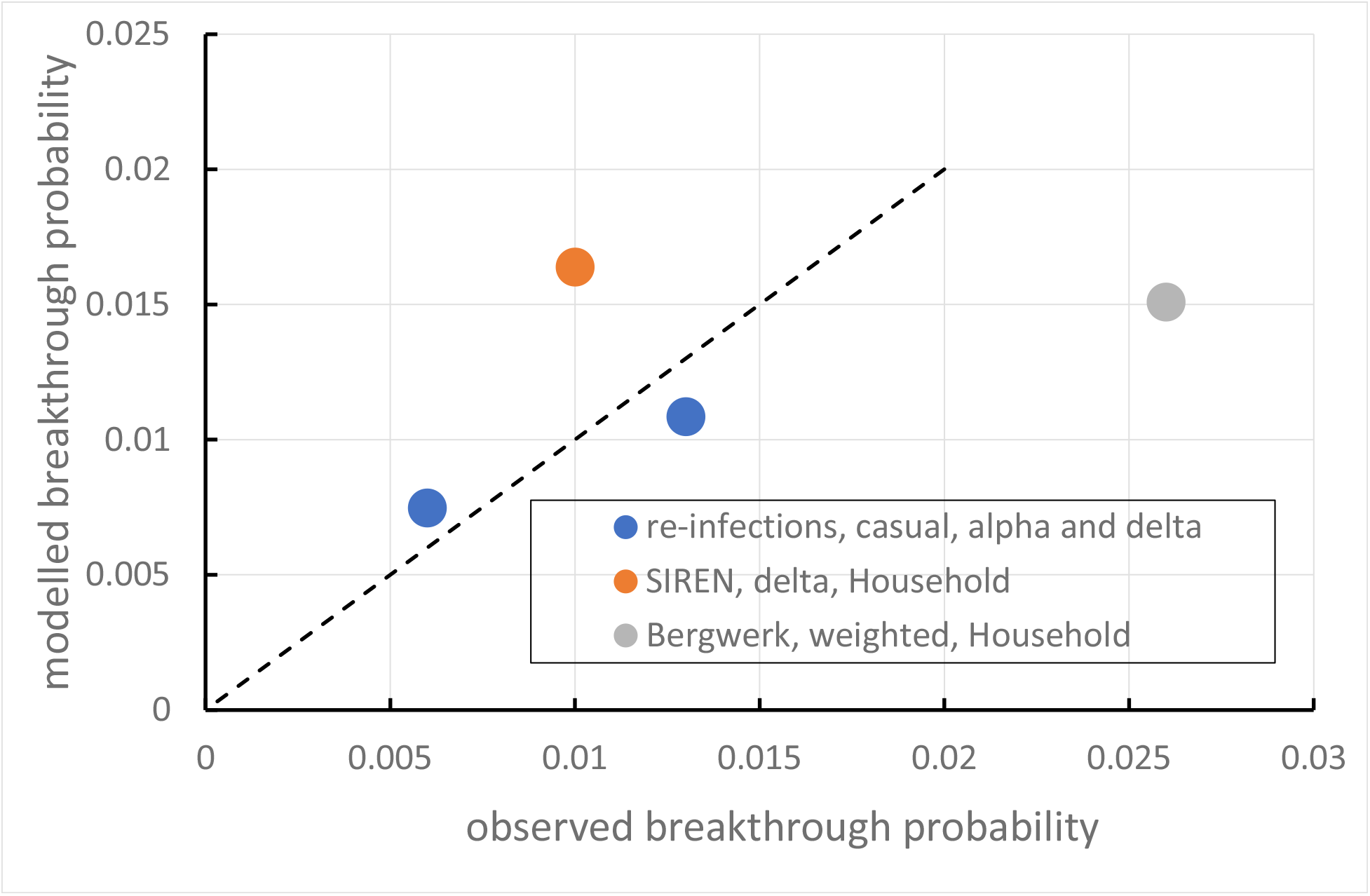
Observed and modelled breakthrough infection probability. P(breakthrough | f_V_(n),E_I_(n), transmission, exposure) = P(breakthrough | f_V_(n), E_I_(n)).P(transmission|exposure).P(exposure) with P(exposure) = 1, P(transmission|exposure) from Table 1 and P(breakthrough | f_V_(n), E_I_(n)) from equation 2(a) with f_V_(n) that for convalescent individuals (reinfections) or Pfizer vaccine (SIREN, Bergwerk^7^) (Figure 2) with σ = 0.44 (pooled data)

### 2. Individual risk based on individual assessment

Individual probability of protection is determined by the high-concentration tail of the antibody concentration distribution. The relative uncertainty in the protection model becomes high at high levels of protection. Figure 4 gives *P(infection*| *n, transmission, exposure)*. Vaccination resulting in an antibody concentration relative to the geometric mean convalescent level, *n* = 1 reduces the probability of infection to approximately 10% of that for an unvaccinated person.

### 3. Individual risk based on a threshold of antibody concentration, T, applied to an individual measurement

Here, the deduced probability of protection is higher than that for risk based on an individual measurement, because it includes the entire population with levels above the threshold. Instead of using equation (3b), equation (3c) is used with *E*_*I*_*(n)* given by equation (2) with the best fit parameters for *k* and *n*_*50*_. The antibody concentration distribution in the incoming population becomes a factor that determines the proportion of incoming people falling into classes defined by the threshold. Figure 6 shows the result, assuming a fully vaccinated incoming population and antibody concentration distribution that for convalescent people. A threshold concentration relative to the geometric mean convalescent level, *n* = 1 reduces the probability of infection to approximately 5% of that for an unvaccinated population. For threshold greater than two times the geometric mean antibody convalescent concentration (corresponding to about 25% of people), the assessed risk does not greatly decrease with further increase of the threshold.

**Figure 6.**
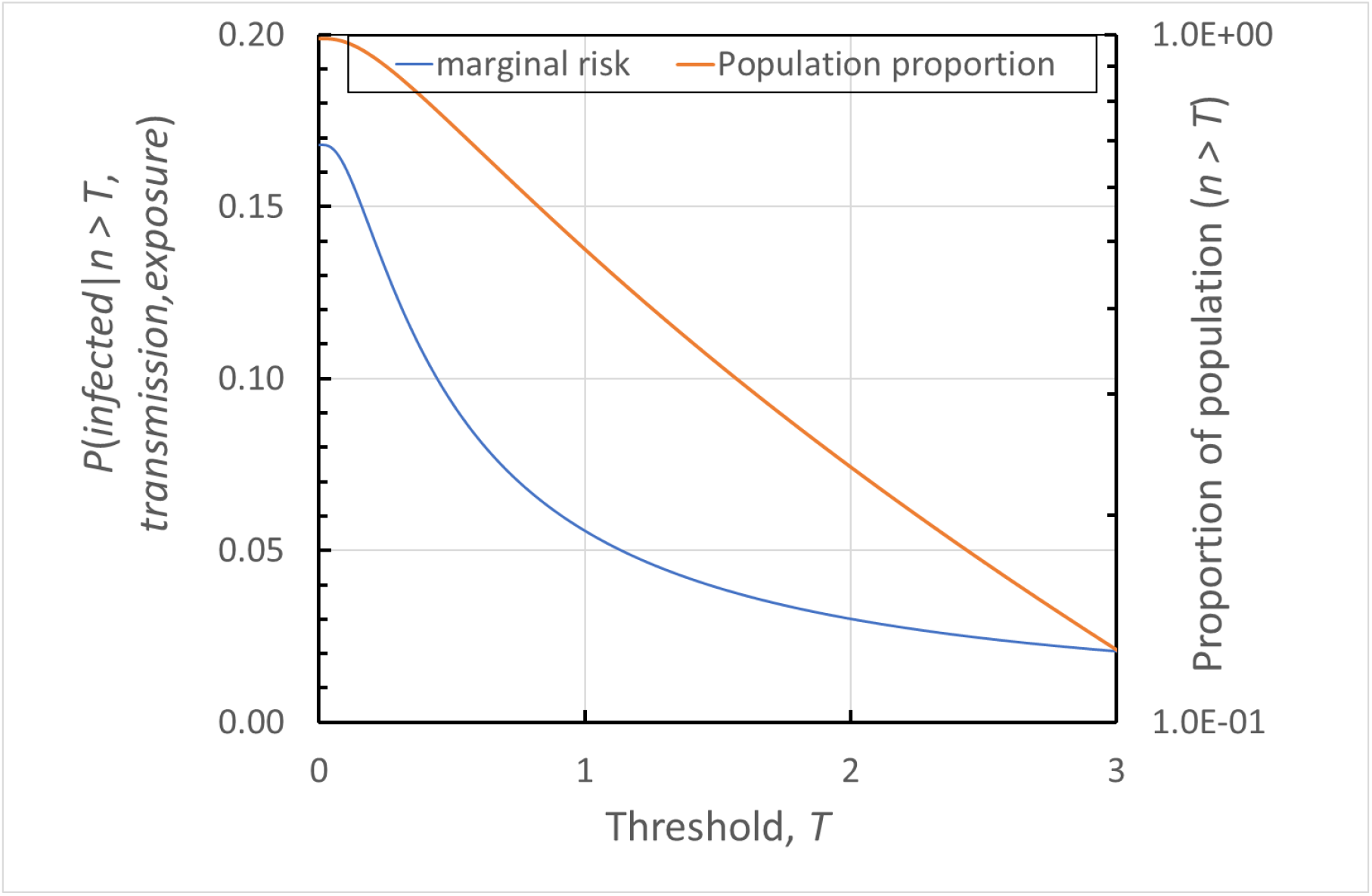
Marginal risk, P(infected| n>T, transmission, exposure) for an incoming vaccinated person given both exposure and transmission, for antibody concentration, n, relative to the geometric mean convalescent concentration, greater than threshold, T, compared with the proportion of the population with n > T (assumes model log-normal convalescent distribution of antibody concentration).

## Discussion: Potential for shortening isolation or quarantine arrangements

### 1. Immunity measurement and viral load measurement are independent assessments of risk

Figure 1 illustrates that sequential measurements of viral load during journey or quarantine are not independent measurements of risk. They assess whether someone has become infected at some time previous to the test. The probability that someone is not infected, given the result of one of these tests, is dependent on the assumptions concerning both the distribution across the infected population and the variation over time following infection of the viral load. The purpose of repeated tests is to identify infected people as quickly as possible in order to minimise the probability of exposure of uninfected people within the travel time or within the time in the managed isolation facility. In contrast, antibody measurement assesses the risk that someone, once exposed, would become infected. Thus the viral load measurement is an assessment of the term in equation (1) *P(transmission*|*exposure)*.*P(exposure)*, evaluated at the time of the measurement. The immunity measurement, being an assessment of probability that is independent of these factors - the probability of infection given all of a particular antibody concentration, transmission and exposure - is a multiplier of the risk assessment as shown in equation 1. Smith *et al*.^4^ and Yang *et al*.^3^ have given model results for importation risk given various testing and quarantine scenarios. The addition of a quantitative immunity measurement applied at one stage at any time during the travel – for example at entry (test 3 in Figure 1) – simply multiplies the assessed risk. Thus for example, for an individual having a measured antibody concentration around the (geometric) mean convalescent level, the assessed risk would be reduced by a factor of approximately 10×.

### 2. Probability of having one or more releases of an infectious person

A way to approach this question is to define a value or a threshold for the individual probability of infection, *P(infection)* in equation 1. Individual measurement of antibody concentration, *n*, together with estimates of *P(transmission*|*exposure)* and *P(exposure)*, assessment of the source country prevalence including an estimate of the fraction of the incoming population who are vaccinated, will define the number of individuals who satisfy this criterion. Smith *et al*.^4^ proposed a classification of source countries based on disease prevalence (cases / 100k population = 0-1, 1-10, 10-100, >100) and then modelled infections / 100k arriving travellers, without border protocols but with the requirement for a pre-flight PCR test (test 1, Figure 1). These assessments of *P(transmission* | *exposure) P(exposure)* were in the range 4×10^-7^ to 2×10^-3^. The values are also consistent with observed positivity rates amongst travellers reported by Yang et al.^3^, increased by a factor of 1.45 to account for the increased infectivity of the delta variant (ratio of secondary attack rates for delta to alpha for non-household contacts) and are consistent with *P(exposure)* based on prevalence rate and *P(transmission* | *exposure)* for the delta variant and non-household contacts. The probability of occurrence of an undetected infection is then, since each of the measures are independent assessments:

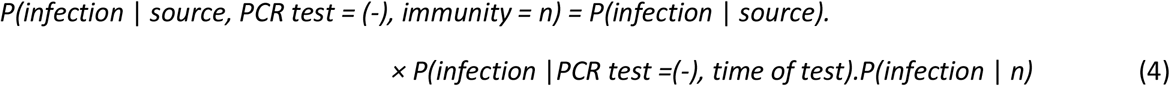

Smith *et al*.^4^ give estimates of the probability of a negative PCR test from an infected person given exposure and time since a previous negative test. These range from 0.3 at 2 days to 0.01 at 15 days. Hence, although the terms *P(infection* |*PCR test =(-), time of test)*.*P(infection* | *n)* are important, the most significant determinant of risk is the source term.

If the probability of observation of an event (in this case an undetected infection) in a sample size *N* is small then the probability of observing one or more events is given by a Poisson distribution:

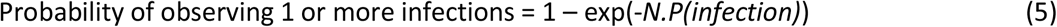

The following section uses equation (5) to compare the risk associated with different border control measures.

### 3. Immunity measurement alone or combined with other controls

The objective is to compare the performance of the current NZ system with that of a system involving accelerated passage through quarantine of people who are identified as vaccinated and with *P(infection*| *n, E*_*I*_*(n), transmission, exposure)* above some defined level, based on an acceptable final risk of having one or more infected persons released into the community and an acceptable proportion of vaccinated people being accelerated. Observational epidemiological study of travellers transiting quarantine in Australia and New Zealand up to 31 March 2021 indicated a failure rate (release of infected cases) of 5.8 / 10^5^ travellers. ^30^ The modelling of Smith et al.^4^ reproduced here indicated a failure rate from high-risk countries for a 14-day quarantine regime of ≈ 2.4 / 10^5^ travellers. The model and observations are reasonably consistent so the modelling of Smith et al is used here.

Taking the threshold for *P(infection*| *n > T, E*_*I*_*(n), transmission, exposure)* = 0.05 would classify approximately half of travellers with convalescent antibody distribution as ‘unsafe’ (Figure 6). For the same risk level, based on the data from Khouri *et al*.^10^, a higher fraction of people vaccinated with AZ and a significantly lower fraction vaccinated with Pfizer would be rejected. Figure 7 then shows the result of application of equation (4) and (5). The assumption is that travel would require a negative pre-departure test taken within 72 hr of departure. Time is thus taken from the time of this test. The risk of exposure during travel or of fraud in the pre-departure test is not included. The model of Khouri *et al*. has not been adjusted to account for the delta variant. Cromer *et al*. have indicated how that might be done ^15^.

**Figure 7.**
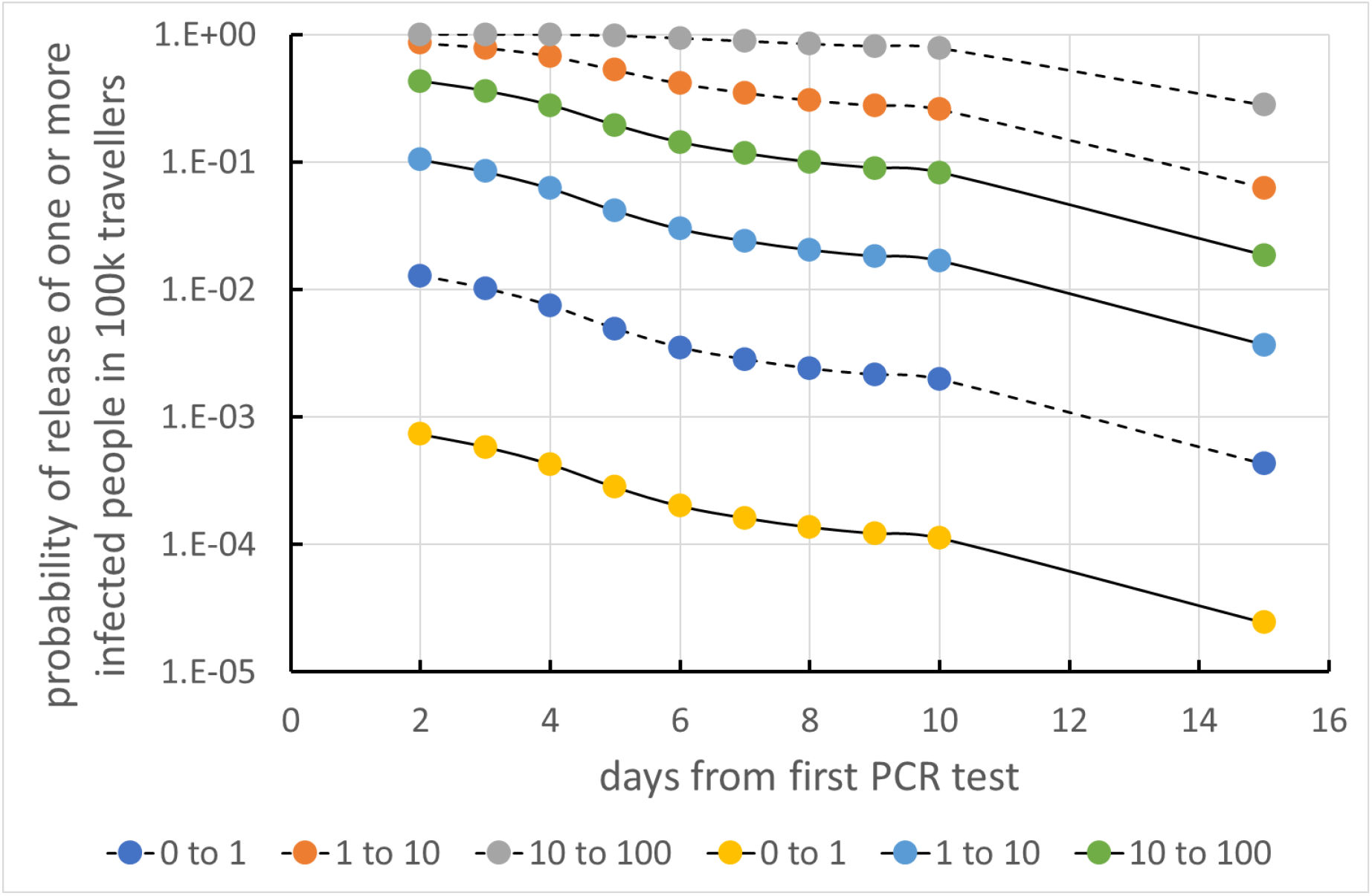
Poisson probability of release into the community of one or more infected travellers, assuming 100,000 travellers, for different source country prevalence, without and with the application of immunity measurement on arrival. Legend: source country prevalence, cases/100,000 population. Dashed lines: without immunity testing. Model of Smith et al.^4^ Solid lines: with immunity testing, assuming a threshold acceptance probability P(infection | n > T) = 0.05.

Figure 7 shows results for different source country prevalence, as noted above. All scenarios assume an on-arrival test and a further test one day before leaving managed isolation. On-departure testing would have a slightly lower probability because of the possibility of fraud.

The use of an on-arrival immunity measurement significantly reduces risk for all scenarios. For travellers from medium-risk countries (prevalence 10-100/100k population) the modelling indicates that use of the immunity measurement gives a risk for 3 days of isolation that is the same or less than that achieved by 14 days of isolation without immunity measurement. For travellers from lower-risk countries, the modelling indicates that use of the immunity measurement would reduce the risk with on-arrival PCR test only and no quarantine to less than that associated with travellers from higher-risk countries with a 14 day isolation without immunity measurement. The uncertainties in the modelling are, however, high. Confirmation through measurement and observation would be needed.

## Data Availability

spreadsheets detailing the calculation available from the author on request

